# Effect of various decontamination procedures on disposable N95 mask integrity and SARS-CoV-2 infectivity

**DOI:** 10.1101/2020.04.11.20062331

**Authors:** Jeffrey S. Smith, Haley Hanseler, John Welle, Rogan Rattray, Mary Campbell, Tacy Brotherton, Tarsem Moudgil, Thomas F. Pack, Keith Wegmann, Shawn Jensen, Justin Jin, Carlo B. Bifulco, Scott A. Prahl, Bernard A. Fox, Nicholas L. Stucky

## Abstract

The COVID-19 pandemic has created a high demand on personal protective equipment, including disposable N95 masks. Given the need for mask reuse, we tested the feasibility of vaporized hydrogen peroxide (VHP), ultraviolet light (UV), and ethanol decontamination strategies on N95 mask integrity and the ability to remove the infectious potential of SARS-CoV-2. FIT test data showed functional degradation by both ethanol and UV decontamination to different degrees. VHP treated masks showed no significant change in function after two treatments. We also report a single SARS-CoV-2 virucidal experiment using Vero E6 cell infection. We hope our data will guide further research for evidenced-based decisions for disposable N95 mask reuse and help protect caregivers from SARS-CoV-2 and other pathogens.

## Introduction

A novel human coronavirus that is now named severe acute respiratory syndrome coronavirus 2 (SARS-CoV-2) emerged from Wuhan, China in December of 2019 (1) and quickly resulted in a global pandemic. The rapid spread of SARS-CoV-2 has created a high demand on personal protective equipment (PPE) and many hospitals worldwide are facing severe shortages. As transmission of SARS-CoV-2 occurs primarily through respiratory droplets, procedure masks and disposable N95 masks in particular have faced severe supply shortages. Contrary to manufacturer recommendations, this unprecedented pandemic has required reuse of these masks. Indeed, many frontline health care workers have adopted individualized mask decontamination strategies with unclear effects on mask integrity and on SARS-CoV-2 decontamination efficacy. Due to more limited supply, more stringent production requirements, and requirement for critical lifesaving aerosol generating procedures N95 masks have become a priority in our health system. This team was tasked with determining feasibility of mask decontamination. Prior studies have investigated how decontamination procedures, including ethanol, ultraviolet light, and vaporized hydrogen peroxide (VHP) alter N95 mask integrity (2–8), but it is unclear how effective these sterilization procedures are at destroying SARS-CoV-2. Here we investigate the effect of different decontamination methods on disposable N95 mask integrity and on eliminating the infectious potential of SARS-CoV-2.

## Results

We first investigated if decontamination strategies, 70% ethanol, ultraviolet light, or VHP, affected N95 mask integrity. We assessed N95 mask integrity through quantitative respirator fit testing (Figure 1A). Quantitative fit testing measures particle concentration inside and outside the respirator and calculates a “FIT score”, the ratio of the two measurements. A FIT score of ³100 is considered sufficient protection from aerosolized particles. Both repeated 70% ethanol exposure and extended exposure to ultraviolet light significantly impaired mask integrity as assessed by FIT scores, consistent with prior reports, although the average score remained above an acceptable functional threshold of 100 in both conditions (Figure 1B,C). VHP maintained an average FIT score of ≥100 with minimal, non-statistically significant degradation of mask components (Figure 1D). A single treatment of 70% ethanol noticeably impaired mask function, even when masks felt dry to the touch (Figure 1C). N95 mask integrity was more greatly impaired at 30 minutes than 4 hours (Supplemental Figure 1). Results were consistent across N95 mask subtypes for both repeated ethanol exposure (Supplemental Figure 2), high-intensity ethanol exposure (Supplemental Figure 3), and VHP (Supplemental Figure 4).

**Figure 1:**
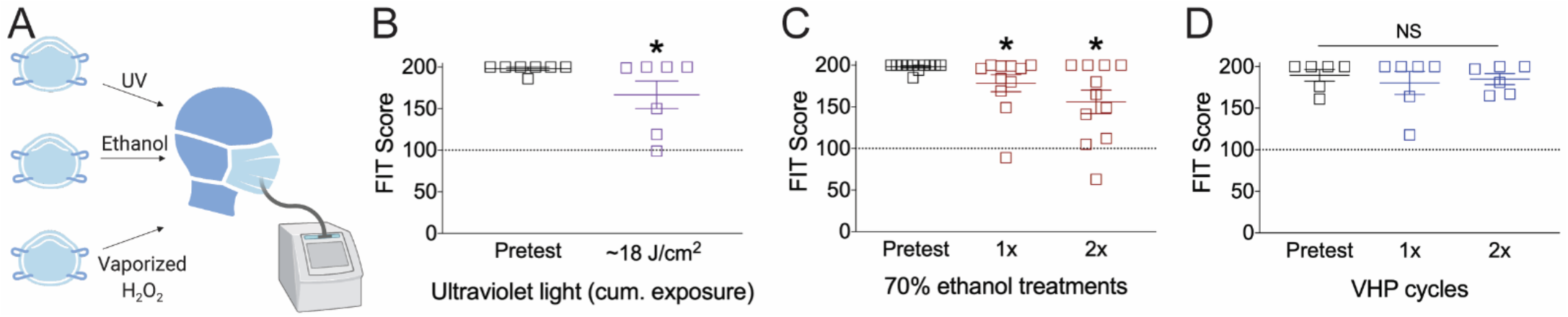
Effect of decontamination methods on N95 mask integrity. **A)** Cartoon of N95 mask decontamination methods. Effect of **B)** UV light, **C)** two applications of 70% ethanol, or **D)** two treatments of vaporized hydrogen peroxide (VHP) on disposable N95 mask integrity. For panel B, *P<0.05, one-tailed t-test; for panels C and D, *P<0.05, one-way ANOVA with Fischer LSD post hoc one-tailed analysis relative to pretest condition. Dashed line at 100 indicates an acceptable FIT score. NS, not significant.

We next tested if decontamination with 70% ethanol, ultraviolet light, or VHP changed viral RNA levels or viral infectivity. In this single experiment, six patients within our hospital system with the highest SARS-CoV-2 titer obtained by nasopharyngeal swab (as assessed by qPCR cycle threshold) were pooled and applied to portions of 1860, 1870+, or 8511 disposable N95 masks and straps, except in the case of the negative control where no virus was applied (Figure 2A). N95 masks subsequently underwent decontamination, aside from the positive controls that were set aside. Following decontamination, N95 masks were immersed in ~3 mL of cell culture media. The media was then sterile filtered, and residual SARS-CoV-2 RNA assessed by RT-qPCR with five different primer sets selective for SARS-CoV-2. SARS-CoV-2 RNA was detected on all masks exposed to virus (Figure 2B, Table 1). However, as the presence of viral RNA does not necessarily indicate viable virus, we then tested the infectious potential of any remaining viable virus exposed to each decontamination condition by applying a fraction of remaining media to Vero E6 cells. We proceeded to culture the virus with cells for 4 days before extracting media to test for the presence of SARS-CoV-2 by both RT-qPCR (Figure 2C, Table 1) and semi-quantitative viral induced cytopathic effects (Supplemental Figure 5).

**Figure 2:**
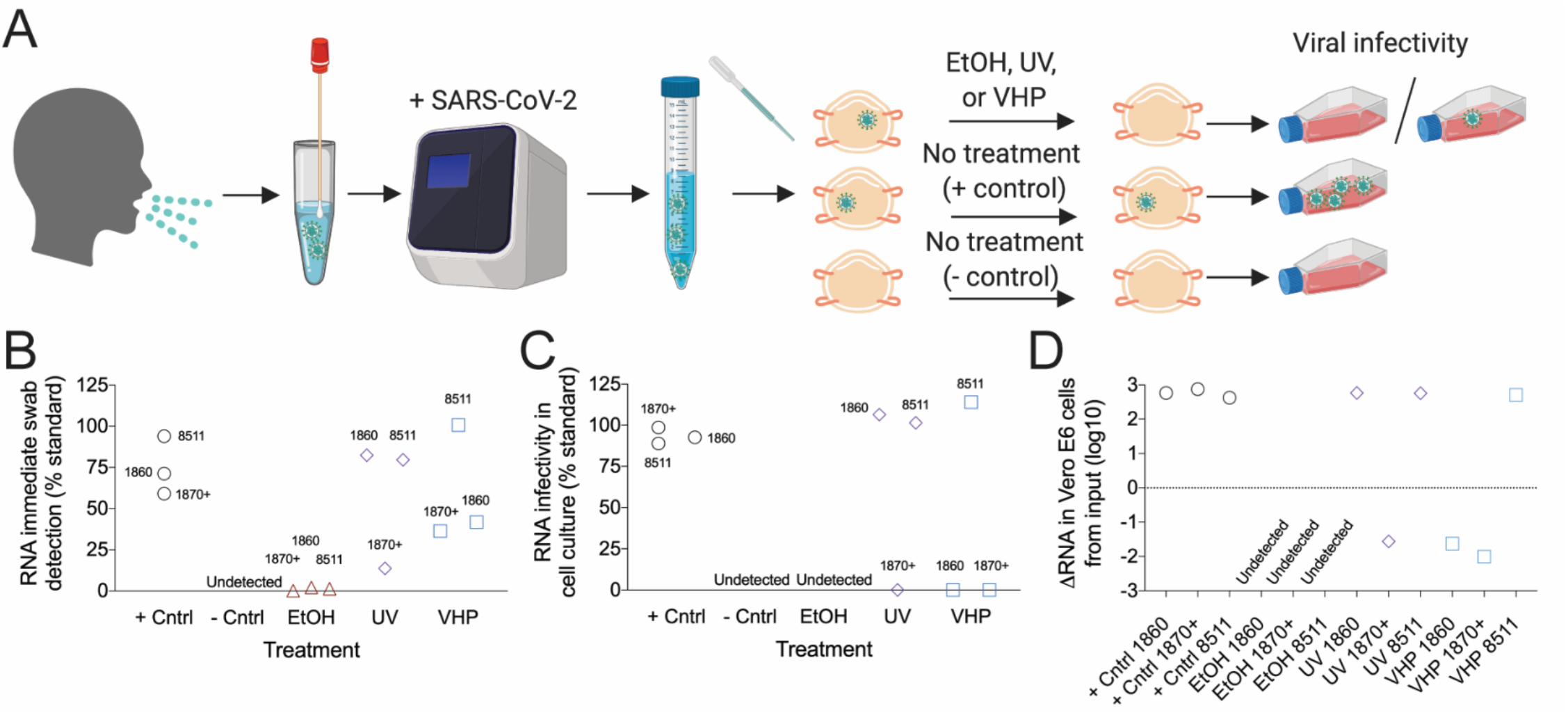
Effect of decontamination methods on SARS-CoV-2 infectivity. **A)** Cartoon of SARS-CoV-2 decontamination experimental design. **B)** RNA detected on the surface of N95 masks immediately after the indicated decontamination treatment. **C)** Infectivity of SARS-CoV-2 in Vero E6 cells after masks underwent the indicated decontamination treatment. **D)** Relative Log10 change of RNA isolated from immediate detection and then detected after infectivity of SARS-CoV-2 culture as assessed by cycle threshold. RNA data displayed is from the SARS-CoV-2 envelope primer set. Data from other primer sets, as well as cycle threshold data, are available in table 1. For B, Data are normalized for the starting SARS-CoV-2 inoculum; for C, data are normalized to the inoculum directly placed in Vero E6 cell culture. Results are from a single experiment.

**Table 1:** Cycle threshold for values of five primer sets for each experimental condition. RNase P is used as an indicator in clinical specimens that sufficient human cellular material was collected, as well as an extraction/procedural control, and was included as reference. RNase P was likely present on N95 masks from skin contact during handling.

| Sample | E-gene<br>(Ct) | N-gene<br>(Ct) | CDC N1<br>(Ct) | CDC N2<br>(Ct) | CDC N3<br>(Ct) | RNase P<br>(Extraction<br>Control)<br>(Ct) |
| --- | --- | --- | --- | --- | --- | --- |
| **SARS-CoV-2 initial RNA** |  |  |  |  |  |  |
| REF | 20.67 | 26.67 | 19.1 | 18.45 | 19.33 | 32.1 |
| ETOH 1860 | 26.23 | 32.63 | 25 | 28.1 | 26.1 | 33.2 |
| ETOH 1870+ | 30.33 | 37.01 | 28.4 | 33.1 | 30.4 | 33.5 |
| ETOH 8511 | 26.9 | 32.62 | 25.2 | 28.5 | 26.5 | 33.3 |
| + Cntrl 1860 | 21.16 | 27.28 | 19.5 | 19.9 | 19.7 | 31.3 |
| + Cntrl 1870+ | 21.43 | 27.19 | 19.4 | 19.9 | 19.6 | 30.9 |
| + Cntrl 8511 | 20.76 | 26.72 | 19 | 19.4 | 19.4 | 32.5 |
| UV 1860 | 20.95 | 27.14 | 19.2 | 19.9 | 19.8 | 33.2 |
| UV Cntrl 1870+ | 23.54 | 29.78 | 21.2 | 22.1 | 21.6 | 34.7 |
| UV Cntrl 8511 | 21 | 27.3 | 19.3 | 19.9 | 19.5 | 34.3 |
| VHP 1860 | 21.93 | 28.57 | 20.2 | 20.6 | 20.6 | 32.8 |
| VHP 1870+ | 22.13 | 28.68 | 20.4 | 21 | 20.8 | 31.6 |
| VHP 8511 | 20.66 | 27.01 | 19 | 19.5 | 19.3 | 31.5 |
| - Cntrl | Undetected | Undetected | Undetected | Undetected | Undetected | 36.1 |
| **SARS-CoV-2 cell culture infectivity RNA** |  |  |  |  |  |  |
| REF | 11.85 | 15.96 | 10.7 | 11 | 11.1 | 32.9 |
| ETOH 1860 | Undetected | Undetected | Undetected | Undetected | Undetected | 43.3 |
| ETOH 1870+ | Undetected | Undetected | Undetected | Undetected | Undetected | 41.7 |
| ETOH 8511 | Undetected | Undetected | Undetected | Undetected | Undetected | 40.1 |
| + Cntrl 1860 | 11.96 | 15.72 | 10.8 | 11.1 | 11.2 | 34.4 |
| + Cntrl 1870 | 11.87 | 15.8 | 10.7 | 11.1 | 11.2 | 34.3 |
| + Cntrl 8511 | 12.02 | 16.32 | 10.9 | 11.2 | 11.3 | 35 |
| UV 1860 | 11.76 | 15.83 | 10.6 | 11 | 11.1 | 35.7 |
| UV Cntrl 1870+ | 28.73 | 34.04 | 25.1 | 26.6 | 26.8 | 38.6 |
| UV Cntrl 8511 | 11.83 | 15.88 | 10.7 | 11.1 | 11 | 33.6 |
| VHP 1860 | 27.33 | 33.22 | 24.6 | 26.2 | 26.1 | 39 |
| VHP 1870+ | 28.79 | 33.23 | 25.3 | 25.9 | 25.9 | 38.3 |
| VHP 8511 | 11.66 | 15.75 | 10.7 | 11.1 | 11.1 | 33.6 |
| - Cntrl | Undetected | Undetected | 36.5 | 35.1 | Undetected | 34.5 |

In descriptive analyses, all three N95 mask types in the positive control cell culture infectivity study had substantially lower cycle thresholds (higher amount of virus) than RNA detected immediately after decontamination corresponding to ~ three log-fold increase in SARS-CoV-2 RNA. N95 masks undergoing different decontamination strategies showed variation in RNA levels (Figure 2D). No RNA was detected in cell culture in any of the three masks treated with 70% ethanol. Supporting the selectivity of our primers, no SARSCoV-2 RNA was detected in any negative control sample in the initial media by any of the five primer sets. The two most sensitive primers did detect low amounts of viral RNA (Ct >35) in the infectivity negative control, likely as a result of slight contamination.

## Discussion

In this study we investigated the effect of different decontamination methods on disposable N95 masks for virucidal effect on SARS-CoV-2 and on N95 mask integrity. This study was initiated early in the course of COVID-19 outbreak when supplies of disposable N95 masks at our institution were limited. Without knowing whether it would be possible to procure new masks, healthcare providers like ourselves had to make urgent decisions about how best to decontaminate existing N95 masks with limited data on decontamination strategies targeting SARS-CoV-2. In order to be compatible with reuse, methods of N95 mask SARS-CoV-2 decontamination must remove the viral threat, be harmless to the mask user, and not compromise the integrity of the various mask elements. The decontamination methods utilized, 70% ethanol, ultraviolet light, and vaporized hydrogen peroxide, have previously been demonstrated to be safe for mask users (3-5,9). We found that any ethanol exposure significantly altered mask integrity, as previously reported (7). We also found that the impact of 70% ethanol on mask integrity appears time dependent. In fact, thirty minutes after 70% ethanol application there was even a larger decline in measured integrity, even though the N95 masks felt dry to the touch. Consistent with prior studies, we did observe a decline in SARS-CoV-2 infectivity (as assessed by Vero E6 culture) after certain decontamination strategies.

There are many limitations to this study. First and foremost, experiments to measure SARS-CoV-2 RNA and infectivity were conducted only once. Our project was initiated to inform decision makers about strategies for mask decontamination within a narrow timeframe. Due to limited resources including continued access to BSL3 laboratory space only a single SARS-CoV-2 decontamination experiment was performed. Another limitation is that clear variation exists between N95 mask type and decontamination efficacy. This could be due to technical replicate variation. However, a reasonable hypothesis to test is that N95 masks with a fluid-resistant coating (healthcare grade) relative to the 8511 (non-healthcare grade) results in less viral uptake and are therefore more likely to be effectively decontaminated. More work is needed to understand this relationship between N95 mask material and decontamination efficacy. Notably, we did not test if ethanol, UV, or VHP impaired N95 fluid-resistant coating on healthcare grade masks. Next, FIT testing, which is utilized by our healthcare system to determine N95 integrity, could in some instances underrepresent actual protection. Likewise, it is also possible ethanol or UV may have resulted in internal mask degradation, producing particulate that may have overestimated the negative impact on N95 mask integrity. While we believe our quantitative FIT testing provides a reasonable estimate of N95 mask function that can aid in comparing imperfect decontamination strategies, we did not conduct the plethora of gold-standard National Institute for Occupational Safety and Health assays required for validation. Another limitation is that we did not test how time alone impacts SARS-CoV-2 infectivity. It is entirely possible that no decontamination, and allowing for decay of virus infectivity over time, is a preferable strategy when faced with no ideal options.

Depending on perspective, the high concentration of SARS-CoV-2 initially applied to N95 masks can be considered either a strength or a weakness. Without access to sophisticated droplet or aerosol generating machines and to avoid unnecessary risk, SARS-CoV-2 containing media was applied directly to the mask samples with a pipette. We intended to deliver the highest challenge possible to assess decontamination efficacy. Samples from the 6 highest titer patients in our healthcare system to date were pooled, and 100uL of this concentrated SARS-CoV-2 containing media was directly infiltrated into the N95 masks with the attempt to expose the middle layer. It is hard to imagine a realistic scenario where healthcare workers would face this degree of mask inoculum. Methods able to decontaminate N95 masks under these intense exposure conditions would likely be highly efficacious in actual practice. However, methods that appear less effective in decontaminating SARS-CoV-2 in our experiment, such as UV, would almost certainly be more effective if masks were challenged in a more realistic exposure scenario. Another possible reason that UV treatment appeared virucidal in only one of the masks we tested is that we chose a dose in the lower of the range of those previously shown to be virucidal (10). In one study a dose of 0.5 J/mc2 was less virucidal than a dose of 1 J/cm2 (11). Regarding VHP treatment, while all masks treated with VHP did not completely eliminate infectious SARS-CoV-2 RNA, the two healthcare grade masks did show a ~five log10 reduction in SARS-CoV-2 RNA relative to the positive control. Further work with additional time points is necessary to confirm if this RNA is infectious or not. In comparision, such a log10 reduction in infectivity would exceed the ‘99.97%’ germidical efficacy quoted by some hand santizers and exceeds the three log10 reduction estimated to fully decontaminate a mask in an influenza model (12). As mentioned above further work across multiple instutitions would be necessary to confirm the degree of this germicidal effect. Overall, it is possible our data inadvertently underestimates the decontamination efficacy of some methods.

Reuse of disposable N95 masks after decontamination has not been recommended under typical circumstances (3). New masks are always preferred. Nevertheless, we recognize that health care systems and front-line providers around the world are currently facing unprecedented shortages of protective respiratory devices and must make decisions that are not ideal. The USA Food and Drug Administration recently authorized emergency use of VHP as a mask decontamination method (13), and our results are consistent with others which showed no significant degredation of mask integrity after two cycles of VHP (6,8). We hope our data will help guide evidenced-based decisions and future experiments that will protect health care providers fighting SARS-CoV-2 and other pathogens.

## Materials and methods

### N95 masks

Disposable N95 masks variants tested included 1860, Aura 1870+, and industrial 8511 were manufactured by 3M (St. Paul, Minnesota, USA). K-N95 disposable masks were manufactured by Jiande Chaomei Daily Chemicals (Mainland China). Masks produced by 3M were combined in main text analysis, with subgroup analyses provided within the supplemental figures. No K-N95 masks had a pretest FIT score of ³100 in our hands, and therefore were not included in experiments summarized in the main text.

### Quantitative evaluation of N95 mask integrity

Quantitative respirator fit testing of disposable N95 masks were conducted using a Portacount Pro 8030 (TSI, Minnesota). In brief, N95 masks were cannulated and an external, air-tight tube attached between the Portacount and mask. Particles that entered the Portacount pro passed through a saturator tube, combined with ethanol vapor, and passed through a condenser tube, effectively enlarging the surface area of the particles. Droplets then passed through a laser, scattering light then quantified by a photodetector. This method allows for detection of particles of various sizes and is not limited to larger particulate. The mask operator then completed up to seven different activities wearing the disposable N95 mask, including normal breathing, deep breathing, head side-to-side, head up-and-down, bending over, jogging, and speaking. Not significant pretest variability existed between users of 3M masks (Supplemental Figure 6). A relative ratio of ambient air particle counts to intramask particle counts was taken, with a ceiling ratio of 200 for each activity. Since few particles should penetrate the high efficiency filter of functional disposable N95 masks, any particles found inside the respirator were conservatively attributed to either face seal leakage or disruption of mask filtration components. An objective ‘fit score’ was then calculated by the formula FIT score = activities/[(1/FF_1)_+(1/FF_2_)…1/FF_7_). Scores of 100 or greater (two log10 reduction in particulate) is considered sufficient for provider projection. Assessments of masks were controlled both intramask and intraoperator, unless otherwise noted.

### 70% Ethanol

70% ethanol obtained by mixing ethanol with 30% deionized water. For disposable N95 mask integrity assessments, 70% ethanol was sprayed 10 times on the mask exterior, the mask was flipped, and sprayed an additional 5 times on the interior, similar to the Italian protocol (14). For overnight application, masks were saturated with 70% ethanol, placed in a sealed plastic bag overnight, removed in the morning and allowed to airdry for ~8 hours prior to FIT testing.

### Vaporized hydrogen peroxide (VHP)

Masks were decontaminated with 30% vaporized hydrogen peroxide using a protocol similar to that previously described (6). Briefly, disposable N95 masks were placed on a metal rack, exterior surface facing upwards. A Bioquell Z vaporizer (Andover, United Kingdom) utilizing 30% hydrogen peroxide solution (Sigma Aldrich, St. Louis, USA) was programmed to gas for 20 minutes at 10 grams per minute, reaching ~500 peak parts per million. Dwell at 4 grams per minute ran for 60 minutes maintain ~420 ppm throughout the full 60 mins. Aeration ran for 210 minutes until reaching safe entrance levels of 1ppm or less. Ambient room temperatures ranged from approximately 22°C ambient to 26°C, and the vaporizer component maintained 120°C during both the gas and dwell phases. Relative room humidity ranged from 38% to 99.5%.

### Ultraviolet light

N95 masks were placed in biosafety cabinet with exterior surface facing towards the UV-C (General Electric 30W Germicidal T8 bulb emitting primarily at 254nm) light source and then flipped to face the interior surface toward the UV source to treat both sides. UV power was measured with a PowerMax-USB PS19 Power Sensor (Coherent Inc, Santa Clara CA) with and without a Schott WG305 filter at the site of mask placement. The difference in the measurements was the UV-C irradiance. Total UV dose was calculated using sensor surface area and time by the equation: irradiance x time = UV dose. To assess mask integrity, a UV dose of 18.4 J/cm^2^ (16 hrs) delivered to the exterior surface and 4.6 J/cm^2^ (4 hrs) delivered to interior surface was used to approximate multiple treatments. To assess virucidal efficacy each side of the mask samples were treated with a UV dose of 0.63 J/cm^2^ (33 min).

### Vero cells and cell culture

Vero cells were obtained from Drs. Victor DeFilippis and Hans-Peter Raue, Vaccine and Gene Therapy Institute, Oregon National Primate Research Center, OHSU. Vero cells were cultured in DMEM 1X, CAT# 1960–044, Lot# 2120580 (Gibco) with 4500 mg/L D-glucose (Gibco) with 10% FBS Cat# PS-500A, Lot# 31C141 (PEAK Serum) 2mM L-glutamine (Lonza) and 50 ug/ml gentamicin sulfate (Lonza). Cells were incubated at 37°C in a humidified incubator with 5% CO_2_. Cells were split 2 to 4 days prior to use and 10^5^ cells were plated into a T25 flask. All work with virus was performed in a certified biosafety cabinet in a BSL2+ negative pressure suite within the Earle A. Chiles Research Institute, Providence Cancer institute or the BSL3 suite of the Regional Pathology Laboratory, Providence St. Joseph Health, Portland, Oregon.

### SARS-CoV-2 detection

After decontamination, N95 masks were immersed in ~3 mL of cell culture media gently agitated for 5 minutes. The media was then sterile filtered, and 600uL was transferred to flasks of Vero E6 cells for infectivity. The remaining approximately 400uL of sample was used for an estimate of SARS-CoV-2 RNA remaining after decontamination. Specific real-time reverse transcriptase–polymerase chain reaction (RT-PCR) targeting the nucleocapsid protein, envelope protein, and RNA-dependent RNA polymerase was used to detect the presence of SARS-CoV-2 similar to that previously described (15). 5 primer sets were used; primers previously validated to target the SARS-CoV-2 envelope and RNA-dependent RNA polymerase (16), as well as the three Centers for Disease Control primers (N1, N2, N3) targeting the SARS-CoV-2 nucleocapsid protein. Nucleic acid extraction was performed using QIAsymphony DSP Virus/Pathogen Mini Kits (Qiagen). qPCR was performed and analyzed on a Roche cobas z480 analyzer or Applied Biosystems 7500 Fast Real-Time PCR Instrument. Equine Arteritis Virus (EAV) and RNase P (shown in table 1) were utilized as an RNA extraction control.

### SARS-CoV-2 viability

Virus viability was assessed by end-point titration in Vero E6 cells similar to that previously described (15). Briefly, a nasopharyngeal swab was collected and placed in viral culture media. Positive SARS-CoV-2 samples were identified as above, and remaining SARS-CoV-2 positive samples with a low cycle threshold (high viral titer) were unthawed and pooled. SARS-CoV-2 positive viral media was transferred on dry ice to appropriate BSL conditions, dethawed, and combined with a saline/albumin mixture that roughly approximated the protein composition of human saliva. 100uL of this SARS-CoV-2 saliva-like solution was then applied to ~1” × 0.5” strips of 1860, 1870+, or 8511 disposable N95 masks, with filter components held together by a single staple. Gentle pressure was applied to increase SARS-CoV-2 saliva-like solution absorbance, and the sample was allowed to rest for 5 minutes. The positive control was transferred to a standard 50mL sterile falcon conical and submerged in DMEM culture media. Negative controls (N95 mask strips without application of SARS-CoV-2 saliva-like solution) were treated similarly. Samples undergoing sterilization were treated similarly as above; pipette application to cover entirety of both surfaces with 70% ethanol, ultraviolet like for 30 minutes per side, VHP, or set aside for 4hrs (positive control). Conical tubes were gently agitated intermittently for 5 minutes. Samples were then subsequently transferred to Vero E6 cells via a 0.2 micron sterile filtration to test for infectivity. 600 µL of sample fluid was transferred to a total of 7 mL of media so all samples were diluted approximately 1:12.

### Specimens

The Regional Providence Institutional Review Board approved the request to obtain leftover viral transport media from nasopharyngeal swabs found to be positive for SARS-CoV-2.

## Data Availability

All data will be available upon reasonable request.

## Acknowledgements

The authors thank Elizabeth Wurdinger, Debra Rhodes and Heather Heidgerken for assistance with N95 FIT testing; Dr. Brian Kendall, Dr. Meera Jain, Dr. Cody Nelson, and Dr. David Martinez for thoughtful discussion. The findings and conclusions in this report are those of the author(s) and do not necessarily represent the official position of Providence Health System or Axovant. Names of specific vendors, manufacturers, or products are included for public health and informational purposes; inclusion does not imply endorsement of the vendors, manufacturers, or products by the authors or by Providence Health System or Axovant.

---

**Supplemental Figure S1:**
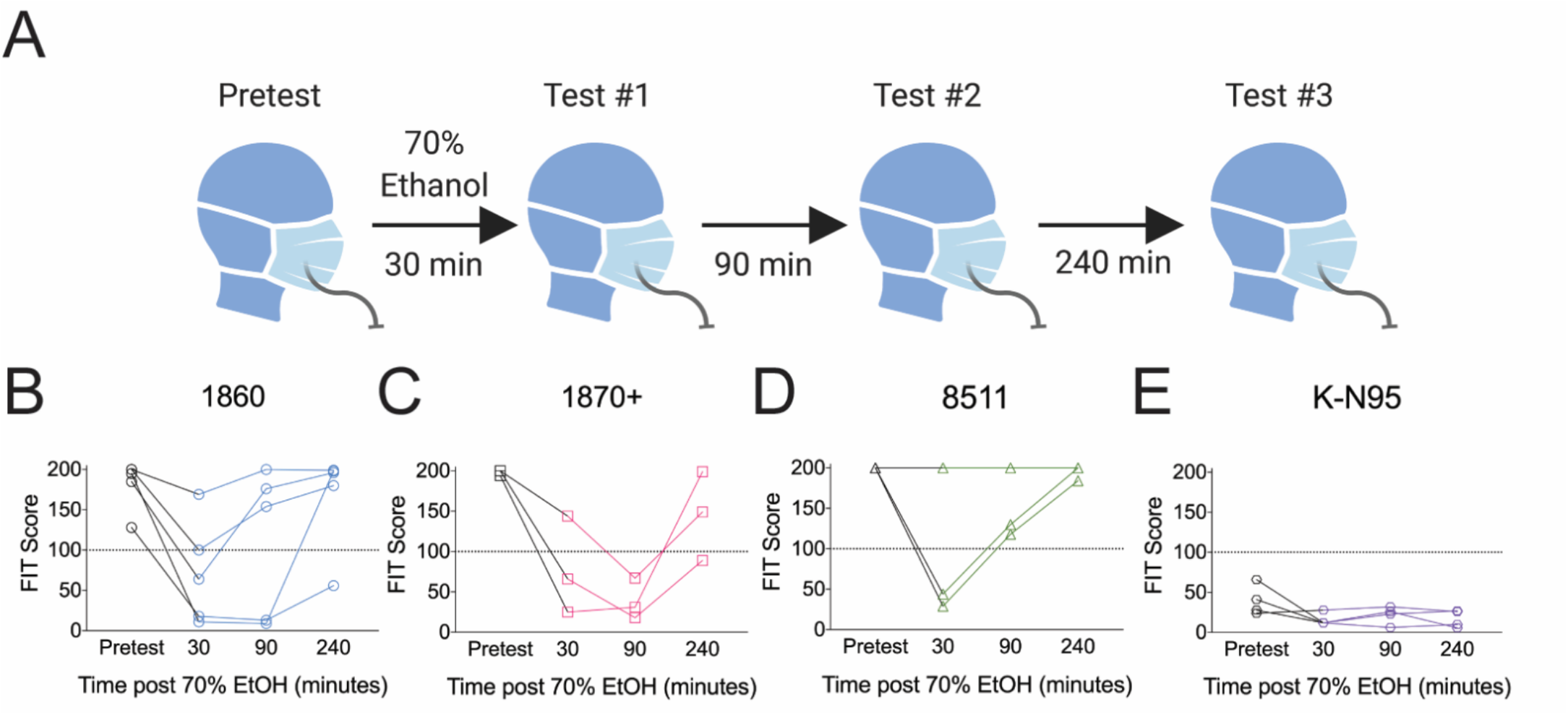
Effect of 70% ethanol treatment on N95 mask integrity is time-dependent. **A)** Cartoon of ethanol treatment time course analyses. **B)** 1860, **C)** 1870+, **D)** 8511, or **E)** K-N95 masks were pretested, treated with 70% ethanol, and re-tested at the indicated time points. Individual N95 masks are shown. No K-N95 masks passed our FIT Pretest and were therefore not included in analyses shown in the main text.

**Supplemental Figure S2:**
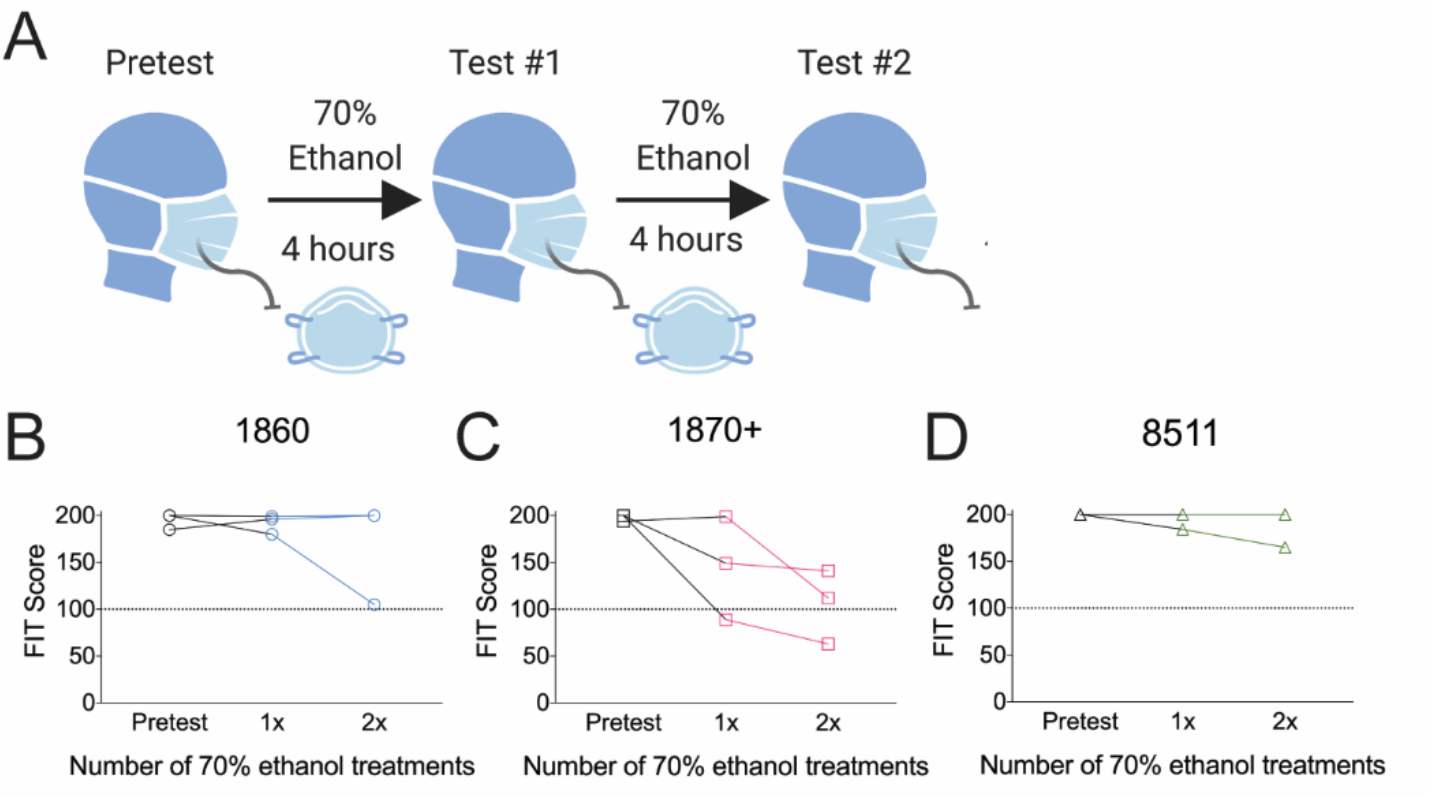
Effect of repeated ethanol on N95 mask integrity. **A)** Cartoon of ethanol treatment time course analyses. **B)** 1860, **C)** 1870+, or **D)** 8511 were pretested, treated the indicated number of times with 70% ethanol, and re-tested. Individual N95 masks are plotted.

**Supplemental Figure S3:**
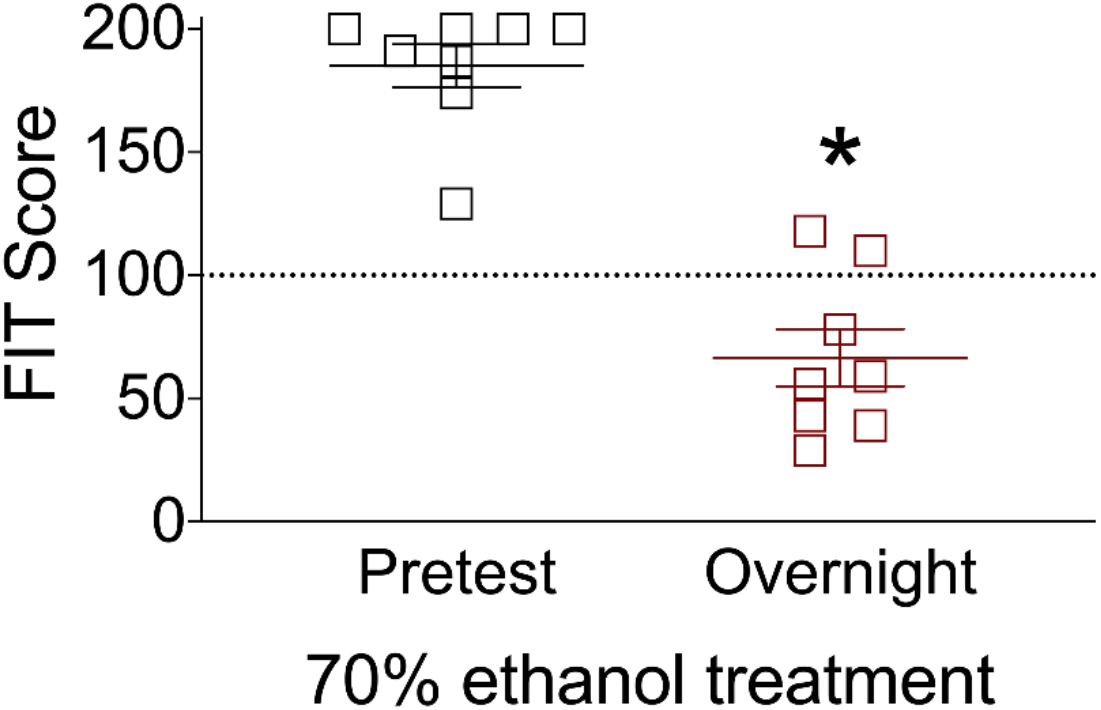
Overnight ethanol treatment impairs N95 mask integrity. After completing FIT pretests, two 1860, three 1870+, and three 8511 disposable N95 masks were saturated with 70% ethanol, sealed within a plastic bag overnight, and allowed to air dry for 8 hours. Masks were then re-tested. Nose guards on some of the 1870+ masks were noted to have detached. *P<0.05, one-tailed t-test. Dashed line at 100 indicates an acceptable FIT score.

**Supplemental Figure S4:**
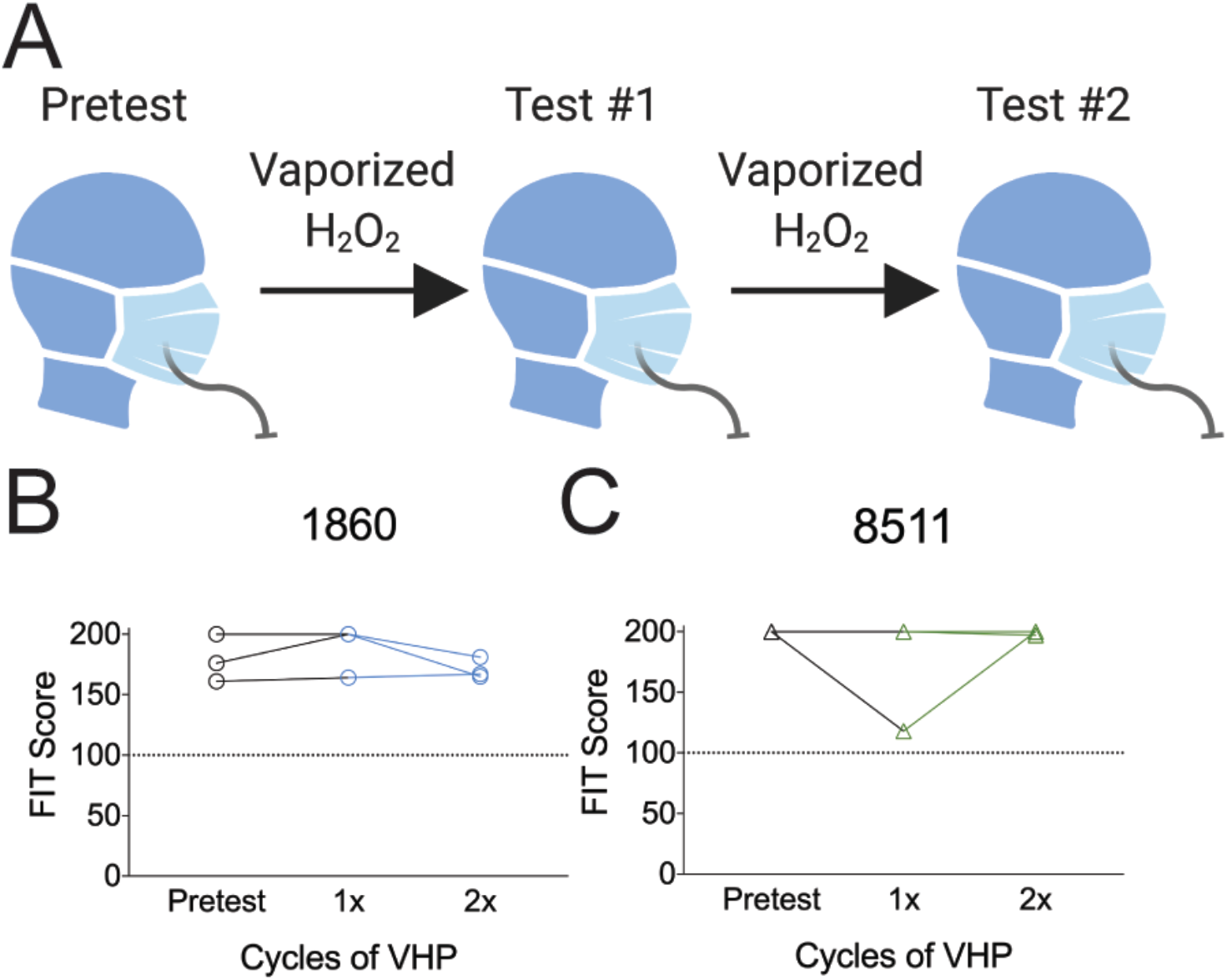
Effect of VHP on N95 mask integrity. **A)** Cartoon of VHP treatment time course analyses. **B)** 1860 or **C)** 8511 N95 masks were pretested, treated the indicated number of cycles of 30% VHP, and re-tested. Individual N95 masks are plotted.

**Supplemental Figure S5:**
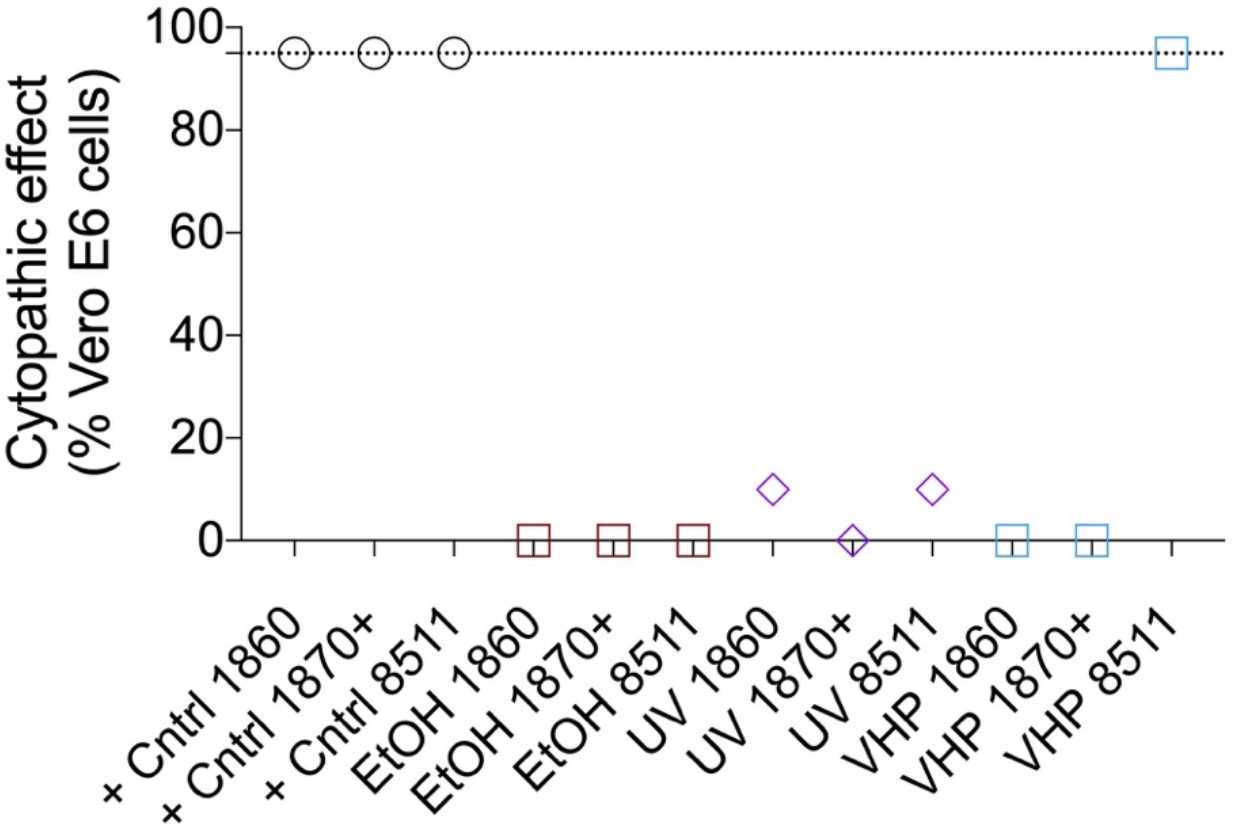
Cytopathic effect (CPE) of SARS-CoV-2 contaminated N95 masks on Vero E6 cells. Vero E6 cells were analyzed under a standard microscope. Cell death in each condition was estimated based on visual inspection and recorded. Dash line represents the CPE of inoculum directly placed in Vero E6 cell culture (positive control).

**Supplemental Figure S6:**
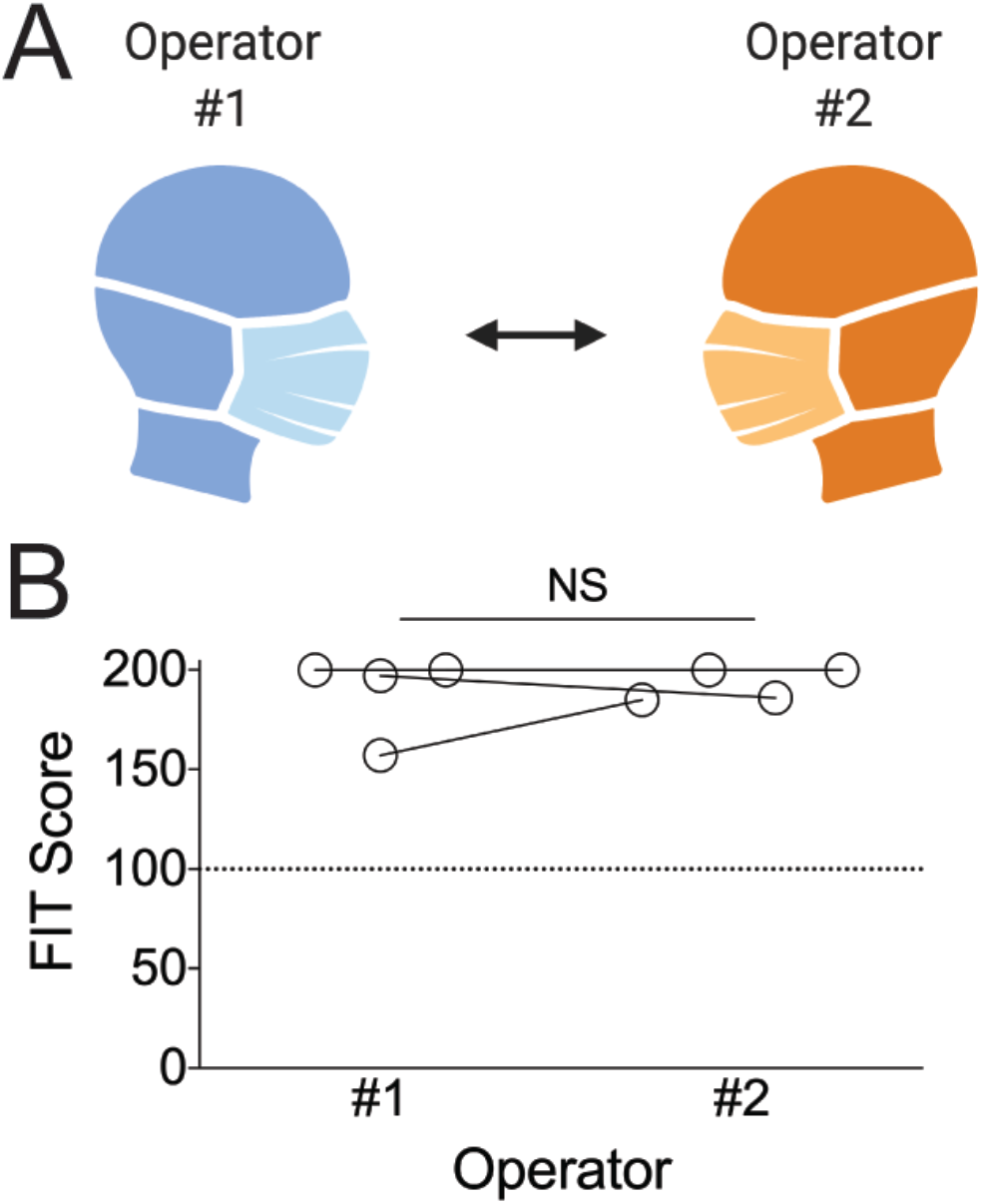
FIT scores are consistent between operators. Two 1860 and two 1870+ masks were analyzed by two different operators. No significant difference in pretest FIT scores were observed. NS, not significant by student’s t-test.

